# Definitions of digital biomarkers: a systematic mapping of the biomedical literature

**DOI:** 10.1101/2023.09.01.23294897

**Authors:** Ana Karen Macias Alonso, Julian Hirt, Tim Woelfle, Perrine Janiaud, Lars G. Hemkens

## Abstract

**Background:** Technological devices such as smartphones, wearables, sensors, or virtual assistants allow to collect data on health and disease processes and are thus increasingly considered a useful digital alternative to conventional biomarkers. We aimed to provide a systematic overview of the emerging literature on “digital biomarkers” with their definitions, features, and citations in biomedical research.

**Methods:** We analyzed all articles in PubMed that used the term “digital biomarker(s)” in title or abstract, considering any study involving humans and any review, editorial, perspective, or other opinion-based article up to 8 March 2023. We systematically extracted characteristics of publications and research studies, and any definitions and features of “digital biomarkers” mentioned. We described the most influential literature on digital biomarkers and their definitions using thematic categorizations of definitions considering the FDA BEST framework (i.e., data type, data collection method, purpose of biomarker), analysing the structural similarity of definitions by performing text analyses (hierarchical clustering on the distance-matrix) and citation analyses (based on citation metrics obtained from OpenAlex via Local Citation Network; last search 26 June 2023).

**Findings:** We identified 415 articles prominently using the term “digital biomarker”. They were published between 2014 and 2023 (median 2021), mostly describing primary research (283 articles; 68%). Most articles did not provide a definition of a digital biomarker (n=287; 69%). The 128 articles providing a definition of a digital biomarker reported 127 different definitions. Of these 127 definitions, 78 considered data collection, 56 data type, 50 the purpose, and 23 were based on all three key components. The 128 articles with a definition were cited a median of 6 times (interquartile range 2-20) with up to 517 citations. Of the ten most frequently cited articles using a definition, all used a different one.

**Interpretation:** The most frequently used definitions for digital biomarkers are highly different and there is no consensus about what this emerging term means. Our overview highlights key defining characteristics of digital biomarkers which can inform the development of a harmonized and more widely accepted definition.

**Funding:** No specific funding.

## INTRODUCTION

Biomarkers are defined as a set of characteristics that are objectively measured and used as indicators of normal biological processes, pathogenic processes, or biological responses that appear due to exposure or therapeutic interventions ^1^. This comprises physiologic, molecular, histologic, and radiographic measurements ^2^. The U. S. Food and Drug Administration (FDA) subclassifies susceptible/risk, diagnostic, monitoring, prognostic, predictive, response, and safety biomarkers ^1^. They highlight that a full biomarker description must include the source or matrix, the measurable characteristic(s), and the methods used to measure the biomarker ^1^.

Digital transformation is continuously influencing activities of daily living and healthcare. This allows digital devices used daily, such as smartphones, wearable devices, sensors, and smart home devices, to provide a new category of biomarkers, often called “digital biomarkers”. In recent years, they became increasingly present in routine care and in research in many areas of medicine, such as cardiology, oncology, or COVID-19. For example, smartphone recorded cough sounds have been used as a digital biomarker to detect asthma and respiratory infections in clinical trials ^3,4^, or deep learning was applied to data from a 3-axis accelerometer to predict sleep/wake patterns ^4,5^. Moreover, such digital biomarkers have spread in the field of neurology, which has a large unmet need for non-invasive and objective biomarkers reflecting cognitive and motor functions that are traditionally assessed with specific tests performed by neurologists ^6^. Beyond monitoring health and disease status, predicting the occurrence and development of diseases would be promising applications of such novel approaches ^7^. Thus, digital biomarkers have the potential to offer valuable insights on the health of patients. They usually have high temporal resolution (up to (quasi-)continuous), are usually objective (and not subject to inter-observer variability), and can have high external validity as they may be applied in the patient’s routine environment (as opposed to e.g. the clinic or a research environment) ^8^.

Many everyday digital tools used mainly for entertainment/leisure purposes (e.g., fitness trackers) are increasingly considered as a source of helpful information that may be transformed into digital biomarkers. Yet, with all this diversity in application and complex interaction with rapidly evolving technology, it becomes necessary to provide a clear and precise definition of the fundamental underlying concepts to facilitate research with and on these novel approaches.

One of the first definitions of this novel type of biomarker was provided by Dorsey et al. in 2017, who defined digital biomarkers as “the use of a biosensor to collect objective data on a biological (e.g., blood glucose, serum sodium), anatomical (e.g., mole size), or physiological (e.g., heart rate, blood pressure) parameter obtained using sensors followed by algorithms to transform these data into interpretable outcome measures, helping to address many of the shortcomings in current measures.” Furthermore, they stated that these new measures “include portable (e.g., smartphones), wearable, and implantable devices, and are by their nature largely independent of raters.” ^9^. A later definition given in 2020 by the European Medicines Agency (EMA) was based upon “digital measures” (“measured through digital tools”) and did not include the requirement of algorithms as a defining feature: “a digital biomarker is an objective, quantifiable measure of physiology and/or behaviour used as an indicator of biological, pathological process or response to an exposure or an intervention that is derived from a digital measure. […]”) ^10^.

Others gave broader definitions including further defining features, for example defining digital biomarkers as “objective, quantifiable, quantitative, physiological, and behavioural data that are collected and measured by means of digital devices such as portables, wearables, implantables, or digestibles. The data collected are used to explain, influence, and/or predict health-related outcomes”^2,6,11^.

Overall, such a disagreement between definitions used by regulators and in articles published in high-impact biomedical journals raised concerns that no clear consensus exists among researchers and users of this novel approach and terminology, increasing the risk for miscommunication and rendering research on digital biomarkers difficult.

We aimed to provide a systematic overview of the emerging literature on digital biomarkers and to characterize the definitions of digital biomarkers that are provided in biomedical journal articles by performing a systematic mapping and citation analysis of all articles that prominently used the term “digital biomarker”.

## METHODS

### Design

We analyzed all articles published at any time in PubMed that prominently used the term “digital biomarker”, i.e., either in title or abstract. We systematically explored definitions of digital biomarkers that are provided and/or referred to in the biomedical literature in a mapping review without a formal assessment of included studies ^12^. We structured our review report to the “Preferred Reporting Items for Systematic Reviews and Meta-Analyses” (PRISMA) guidance, where applicable ^13^. We did not use a pre-specified protocol.

### Eligibility criteria, information source, and search strategy

We searched PubMed and included all articles mentioning “digital biomarker” or “digital biomarkers” in their title or abstract (by searching PubMed for “digital biomarker*[tiab]”; date of last search: March 8, 2023). We excluded animal research.

### Study selection

One reviewer (AKM) screened titles, abstracts, and full texts for eligibility; with confirmation by a second reviewer (JH or LGH), if necessary.

### Data extraction

We developed a spreadsheet to structure the data extraction process. One reviewer (AKM) extracted data with confirmation by a second reviewer (JH or LGH), if necessary.

We extracted from every article: author(s), publication year, title, journal, corresponding author, and country of correspondence, article type (i.e., primary research, review, or other type [e.g., editorial, comment, opinion-based letter]). Of primary research articles, we additionally extracted definitions of digital biomarkers that are provided and/or referred to (based on a semantic search for indicators of definition such as “digital biomarkers are”, “… are defined as”, “… can be defined”, “the definition of … is”), medical context, and whether the article is about the development and/or validation of a digital biomarker. The number of global citations was obtained by using metadata from OpenAlex ^14^; accessed via the Local Citation Network ^15^ (as of June 26, 2023).

### Data analysis and categorization of definition components

We considered the BEST (Biomarkers, EndpointS, and other Tools) framework, developed by the FDA and U.S. National Institutes of Health (NIH) “with the goals of improving communication, aligning expectations, and improving scientific understanding” to derive components of definitions for digital biomarkers^1^. We defined definitions as duplicates when they used the same sequence of words. We categorized identified digital biomarker definitions if they contained information on three key components, i.e. the (i) type of data that is measured (e.g., objective, continuous, quantifiable, or quantitative data), (ii) data collection method (e.g., sensor, computer, portable, wearable, implantable, or digestible), and (iii) purpose of the digital biomarker (e.g., used as measures of disease progression or to explain health related outcomes). We illustrate the frequency of various terminologies used in all provided definitions with a word cloud ^16^. We analysed the structural similarity of definitions that were provided without a reference by performing hierarchical clustering on the distance-matrix containing pairwise “Indel”-distances, i.e. “the minimum number of insertions and deletions required to change one [definition] into the other” ^17^. Since we aimed at exploring how digital biomarkers are defined in the biomedical literature, we did not critically assess the included articles and studies. For the analysis of citations, we calculated the quotient of number of global citations (retrieved by the Local Citation Network ^15^) and years since publication per article. To create a citation network of citing and cited relationships between the articles, we used the Local Citation Network with the OpenAlex scholarly index ^15,18^.

We used descriptive statistics by reporting numbers and percentages. For all analyses, we used R (version 4.2.2) or Python (3.11.4).

## RESULTS

We identified 415 articles that had “digital biomarker” in their title or abstract (Supplementary Material S1). The first article was published in 2014 (median publication year 2021; Figure 1; Supplementary Material S2). Most articles described primary studies (n=283; 68%) and were published in in digital medicine specialty journals, including Digital Biomarkers (n=35; 8%), Journal of Medical Internet Research (n=21; 5%) or NPJ Digital Medicine (n=19; 4%; Table1). Of the 415 articles, 128 (31%) provided at least one definition of a digital biomarker.

**Figure 1.**
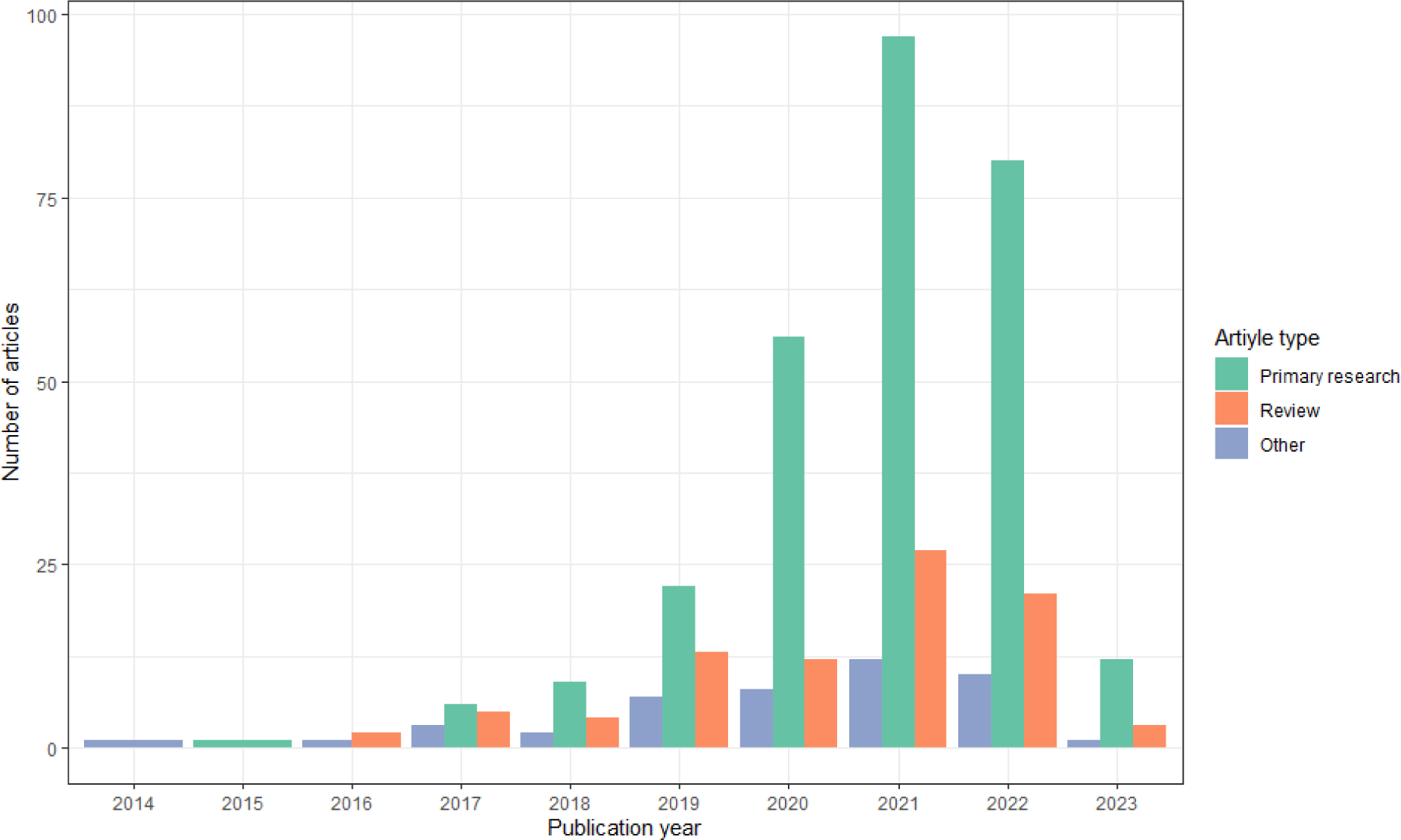
The annual number of published article types referring to Digital Biomarkers as of March 8, 2023 (n=415)

### Characteristics of articles providing a definition of digital biomarker

The 128 articles with a definition of digital biomarker were published between 2015 and 2023 (median: 2021). Of them, 59 articles were primary studies, 50 were reviews, and 19 were other types of articles (Table 1).

**Table 1.**
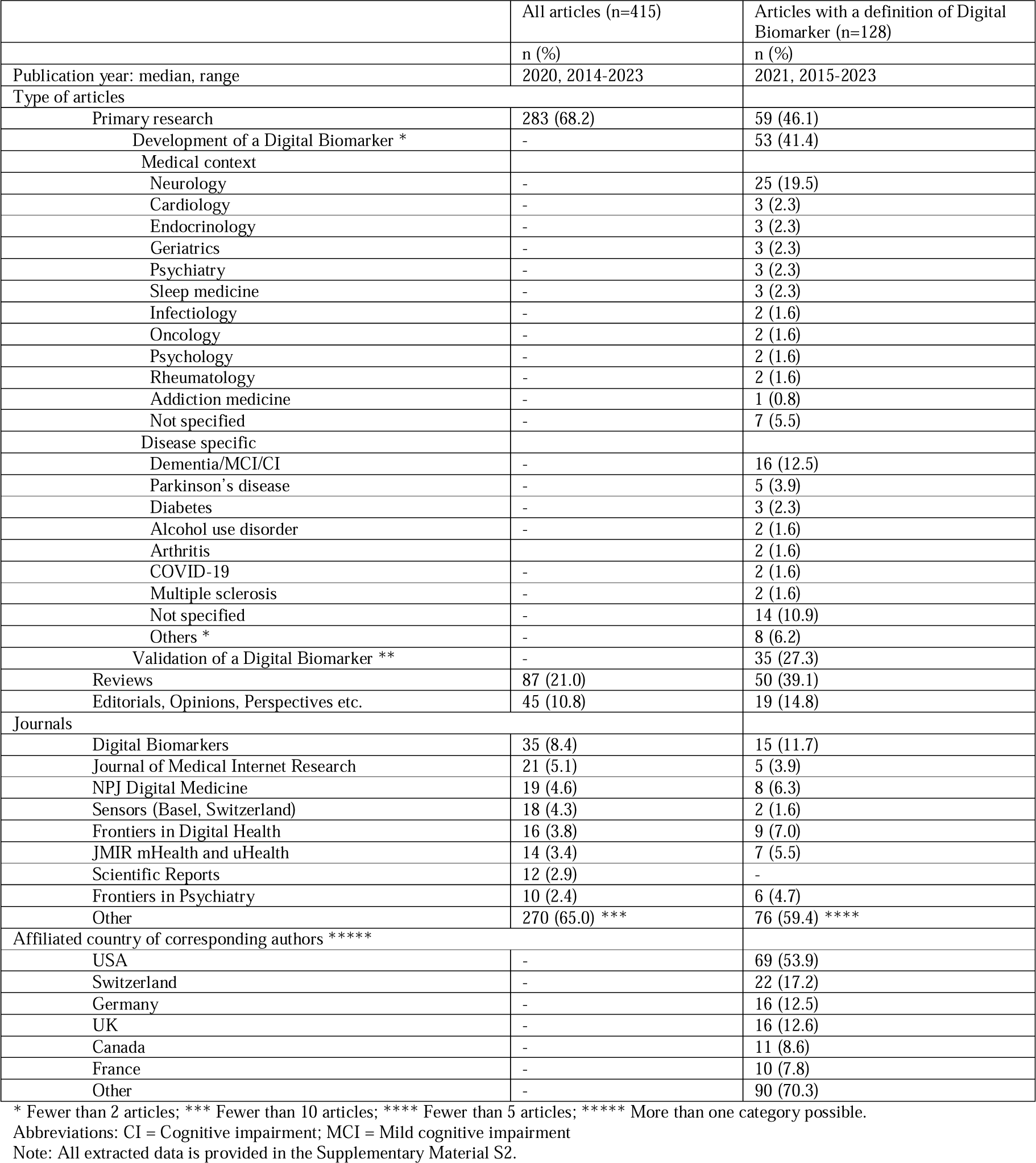
Characteristics of all 415 articles in PubMed using “digital biomarker” in title or abstract.

|  | All articles (n=415) | Articles with a definition of Digital Biomarker (n=128) |
| --- | --- | --- |
|  | n (%) | n (%) |
| Publication year: median, range | 2020, 2014-2023 | 2021, 2015-2023 |
| Type of articles |  |  |
| Primary research | 283 (68.2) | 59 (46.1) |
| Development of a Digital Biomarker * | - | 53 (41.4) |
| Medical context |  |  |
| Neurology | - | 25 (19.5) |
| Cardiology | - | 3 (2.3) |
| Endocrinology | - | 3 (2.3) |
| Geriatrics | - | 3 (2.3) |
| Psychiatry | - | 3 (2.3) |
| Sleep medicine | - | 3 (2.3) |
| Infectiology | - | 2 (1.6) |
| Oncology | - | 2 (1.6) |
| Psychology | - | 2 (1.6) |
| Rheumatology | - | 2 (1.6) |
| Addiction medicine | - | 1 (0.8) |
| Not specified | - | 7 (5.5) |
| Disease specific |  |  |
| Dementia/MCI/CI | - | 16 (12.5) |
| Parkinson's disease | - | 5 (3.9) |
| Diabetes | - | 3 (2.3) |
| Alcohol use disorder | - | 2 (1.6) |
| Arthritis | - | 2 (1.6) |
| COVID-19 | - | 2 (1.6) |
| Multiple sclerosis | - | 2 (1.6) |
| Not specified | - | 14 (10.9) |
| Others * | - | 8 (6.2) |
| Validation of a Digital Biomarker ** | - | 35 (27.3) |
| Reviews | 87 (21.0) | 50 (39.1) |
| Editorials, Opinions, Perspectives etc. | 45 (10.8) | 19 (14.8) |
| Journals |  |  |
| Digital Biomarkers | 35 (8.4) | 15 (11.7) |
| Journal of Medical Internet Research | 21 (5.1) | 5 (3.9) |
| NPJ Digital Medicine | 19 (4.6) | 8 (6.3) |
| Sensors (Basel, Switzerland) | 18 (4.3) | 2 (1.6) |
| Frontiers in Digital Health | 16 (3.8) | 9 (7.0) |
| JMIR mHealth and uHealth | 14 (3.4) | 7 (5.5) |
| Scientific Reports | 12 (2.9) | - |
| Frontiers in Psychiatry | 10 (2.4) | 6 (4.7) |
| Other | 270 (65.0) *** | 76 (59.4) **** |
| Affiliated country of corresponding authors ***** |  |  |
| USA | - | 69 (53.9) |
| Switzerland | - | 22 (17.2) |
| Germany | - | 16 (12.5) |
| UK | - | 16 (12.6) |
| Canada | - | 11 (8.6) |
| France | - | 10 (7.8) |
| Other | - | 90 (70.3) |

\* Fewer than 2 articles; \*\*\* Fewer than 10 articles; \*\*\*\* Fewer than 5 articles; \*\*\*\*\* More than one category possible.
Abbreviations: CI = Cognitive impairment; MCI = Mild cognitive impairment
Note: All extracted data is provided in the Supplementary Material S2.

Almost all primary studies described the development of one or more digital biomarkers (53 of 59 articles), and many described a validation process of biomarkers (35 of 59 articles). The most frequent medical field of the primary research articles that described the development of one or more digital biomarkers was neurology (25 of 53), while the spectrum of medical fields was overall very wide (Table 1). The most frequent diseases were dementia and related disorders (16 of 53 articles, i.e., [mild] cognitive impairment or Alzheimer’s disease), Parkinson’s disease (5 of 53 articles), and diabetes (3 of 53 articles), with numerous other conditions addressed in one or two studies (e.g., atrial fibrillation, cervical cancer, depression, heart failure, and muscular dystrophy; Supplementary Material S2).

The corresponding authors were mostly from the United States of America (69 of 128 articles), Switzerland (22 of 128 articles), Germany (16 of 128 articles), and the United Kingdom (16 of 128 articles; Table 1).

The articles were cited a median of 6 times (range 0-517, interquartile range (IQR) 2-20, overall 2,705); on average 2 times per year (range 0-86, IQR 1-5; Supplementary material S2). We show the citation network (i.e., citing and cited relationships within the sample of these 128 articles) online (https://LocalCitationNetwork.github.io/?fromJSON=Digital-Biomarker-Definitions.json).

### Definitions of digital biomarkers

Overall, 128 articles reported between 1 and 7 definitions (median 1, IQR 1 to 2). In 91 articles, at least one reference was provided for these definitions made by the authors (median 1, range 1-13, IQR 1 to 2, overall 274 references); for 37 articles with 51 definitions, no reference was provided (Supplementary Material S2).

The mostly used references to support the definitions were Coravos, Khozin et al. (2019) ^4^ (referenced by 51 of 91 articles); Dorsey et al. (2017) ^9^ (11 articles); Califf (2018) ^19^ (9 articles); Piau, Wild et al. (2019) ^20^ (9 articles); Babrak et al. (2019) ^6^ (8 articles), and Coravos, Goldsack et al. (2019) ^21^ (8 articles). All these articles were among the 415 articles analysed here. The original definitions in these top-cited articles can be found in Table 2. Other references were used by less than 5 articles.

**Table 2.** The top cited definitions of Digital Biomarkers within the 415 articles.

| Authors (year); Reference | Number of articles citing this definition in the 415 articles | Definition (original quote) |
| --- | --- | --- |
| Coravos, Khozin et al. (2019) <sup>4</sup> | 51 | "We describe an emerging class of biomarker, the "digital biomarker", which has important implications for both clinical trials and clinical care. "Digital" refers to the method of collection as using sensors and computational tools, generally across multiple layers of hardware and software. The measurements are often made outside the physical confines of the clinical environment using home-based connected products including wearable, implantable, and ingestible devices, and sensors. Digital biomarkers span a broad range of diagnostic and prognostic measurements." |
| Dorsey et al. (2017) <sup>9</sup> | 11 | "Digital biomarkers – the use of a biosensor to collect objective data on a biological (e.g., blood glucose, serum sodium), anatomical (e.g., mole size), or physiological (e.g., heart rate, blood pressure) parameter followed by the use of algorithms to transform these data into interpretable outcome measures can help address many of the shortcomings in current measures. These new measures, which include portable (e.g., smartphones), wearable, and implantable devices, are by their nature largely independent of raters. They are, therefore, not prone to rater bias. The goal of digital biomarkers is to maximize the ecological validity and temporal and spatial resolution of capturing motor and nonmotor phenomena that are expected to change over time." |
| Piau, Wild et al. (2019) <sup>20</sup> | 9 | "Digital biomarkers are defined here as objective, quantifiable, physiological, and behavioral data that are collected and measured by means of digital devices, such as embedded environmental sensors, portables, wearables, implantables, or ingestibles. Digital biomarkers allow objective, ecologically valid, long-term follow-up with frequent or continuous assessment that can be minimally obtrusive or function in the background of everyday activity." |
| Babrak et al. (2019) <sup>6</sup> | 8 | "Digital biomarkers are objective, quantifiable, physiological, and behavioral measures that are collected by means of digital devices that are portable, wearable, implantable, or ingestible. These data are often used to explain, influence, and/or predict health-related outcomes. Digital biomarkers fall within the scope of traditional biomarkers in relation to addressing health related questions, with use of a digital and portable technology that adds new dimensions, unique features, and challenges. digital biomarkers are usually less or non-invasive, modular, and often cheaper to measure. They can produce qualitative and quantitative measurements, but most importantly, they provide easier and cheaper access to continuous and longitudinal measurements." |
| Califf (2018) <sup>19</sup> | 8 | "... digital biomarkers derived from sensors and mobile technologies. ...these data are in large part derived from new sources including smartphones and wearable electronic devices and facilitated by novel technologies that allow for the streaming and storage of complex data, standards for evaluating these biomarkers are just now developing." |
| Coravos, Goldsack et al. (2019) <sup>21</sup> | 8 | "A digital biomarker could be any of the seven BEST biomarker types. The term digital refers to the method of collection as using sensors and computational tools, generally across multiple layers (e.g., a full stack) of hardware and software." |

In total, the 128 articles reported 202 definitions; 75 of which were duplicates. Hence, we identified 127 unique definitions across the 128 articles.

The ten most frequently used terms that most of the 127 unique definitions contained were “digital” (125 of 127 definitions; 98%), “biomarkers” (109 of 127 definitions; 85%), “data” (62 of 127 definitions; 48%), “collected” (55 of 127 definitions; 43%), “devices” (50 of 127 definitions; 39%), “health” (42 of 127 definitions; 33%), “physiological” (37 of 127 definitions; 29%), “objective” (37 of 127 definitions; 29%), “wearable” (34 of 127 definitions; 26%), and “behavioral” (33 from 127 definitions; 25%; Figure 2).

**Figure 2.**
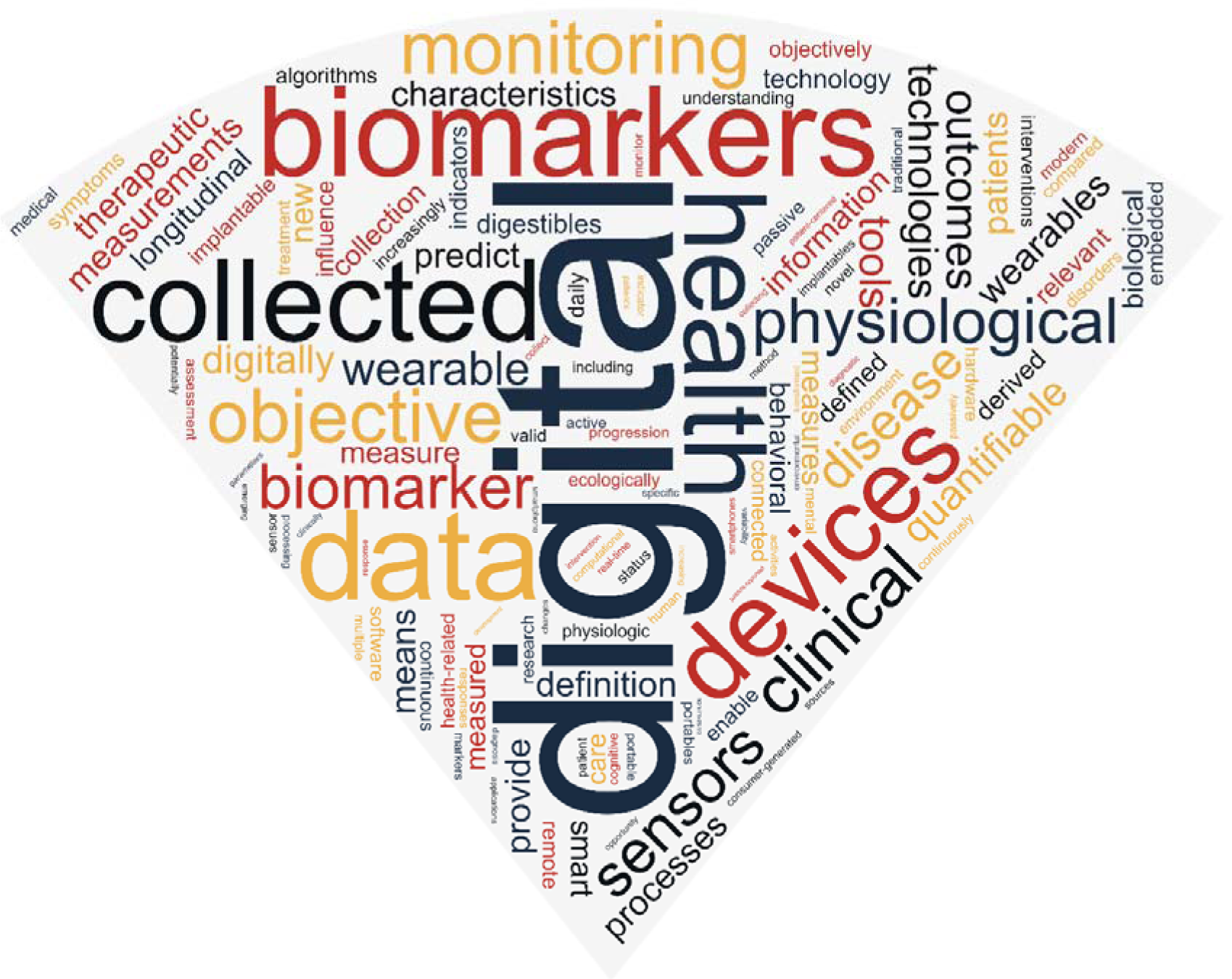
Word cloud with the most frequently used terms in the analysed digital biomarker(s) definitions.

Of the 127 unique definitions, 56 definitions refer to the type of data that are collected, 78 definitions contain information on the data collection method, and 50 definitions provide information on the purpose of the digital biomarker. Only 23 of 127 definitions involve all three components and 26 contain none of these components (Table 3; Supplementary Material S3; Supplementary Material S2). There were almost no structural similarities between the 51 identified definitions in 37 articles without a reference (for those with a reference, similarities such as paraphrasing are expected); Supplementary material S3.

**Table 3.** The definitions of digital biomarkers that considered the type of data, data collection method, and purpose of a digital biomarker (n=23)

| Authors (year), Reference | Definition (original quote) |
| --- | --- |
| Andrade et al. (2022) <sup>25</sup> | "Digital biomarkers may have a place as an objective, accurate, and low-cost patient metric to support risk stratification and clinical planning. Digital biomarkers use digital information to objectively measure biological and pathological processes and have the potential to overcome some of the above-mentioned limitations of conventional prognostic tools. Digital data, in particular data from accelerometers and other wearable sensors, are a non-invasive, passively collected low-cost source of individual information. Further exploration of clinical uses for these data may improve clinical decision-making with minimal risk and cost." |
| Bartolome et al. (2022) <sup>26</sup> | "Digital biomarkers refer to objective, quantifiable physiological, and behavioral measures that are collected by means of digital devices, such as wearable devices, for the purpose of outcomes explaining, influencing, or predicting health. However, unlike traditional biomarkers that provide a "snapshot view" based on limited measurements collected over time, digital biomarkers are often derived from longitudinal and continuous measurements, and thus can capture dynamic changes in health and related outcomes." |
| Bijlani et al. (2022), Nam et al. (2019), Parziale et al. (2022), Phillips et al. (2017), and Wright et al. (2018) <sup>27-31</sup> | "Digital biomarkers are consumer-generated physiological and behavioral measures collected through connected digital tools that can be used to explain, influence and/or predict health-related outcomes. Health-related outcomes can vary from explaining disease to predicting drug response to influencing fitness behaviors. In our definition of digital biomarkers, we exclude patient-reported measures (e.g., survey data), genetic information, and data collected through traditional medical devices and equipment. These data types, though still a key component of research and clinical care that may be stored digitally, are not digitally measured or truly dependent on software." |
| Harms et al. (2022) <sup>32</sup> | "Digital biomarkers are defined as objective, quantifiable physiological and behavioral data that are collected and measured by means of digital devices. Their use has revolutionized clinical research by enabling high-frequency, longitudinal, and sensitive measurements. Digital biomarkers are that the latter are collected via digital devices and can be collected outside of traditional clinical settings. The digital devices collecting these biomarkers can include wearables, implantables, ingestible devices, and smartphones and tablets. Examples of digital biomarkers are objective consumer-grade data such as voice, temperature, activity, gait, blood oxygen, heart rate, touch, and augmented reality, all collected via mobile and wearable technologies. As opposed to standard clinical measures, digital biomarkers enable high-frequency, longitudinal, and objective measurements, largely independent of the clinical rater. Digital biomarkers can continuously monitor patients to assess therapy response and disease progression without the need for clinical assessment. Moreover, they often exhibit higher sensitivity than traditional clinically used methods, enabling early predictive diagnostics by identifying patients at risk of overt clinical disease." |
| Katsaros et al. (2022) <sup>33</sup> | "Digital biomarkers are objective measurements of physiological, pathologic, or anatomic characteristics continuously collected outside the clinical environment via home-based connected devices. Passively collecting data from patients' mobile or wearable devices potentially offers a convenient and unobtrusive method to prospectively identify psychosocial burden and deliver tailored social support to the right patients at the right time." |
| Motahari-Nezhad et al. (2022) <sup>34</sup> | "Sensors and digital devices have revolutionized the measurement, collection, and storage of behavioral and physiological data, leading to the new term digital biomarkers. Digital biomarkers are measured across multiple layers of the hardware (eg, sensors) and software of medical devices that capture signals (behavioral and physiological data) from patients. Digital biomarkers can increase diagnostic and therapeutic precision in the modern health care system by remotely and continuously measuring reliable clinical data and allowing continuous monitoring and evaluation. Captured by wearable, implantable, and ingestible devices and sensors, digital biomarkers can be used at home to provide clinical data, collecting data that is not possible in the clinical setting. This information can improve physicians' and patients' decisions, personalize the treatment, and predict diseases' current and future status." |
| Shandhi et al. (2022) <sup>35</sup> | "Multiple studies suggest the utility of digital biomarkers, objective and quantifiable digitally collected physiological and behavioral data (e.g., resting heart rate (RHR), step count, sleep duration, and respiratory rate), collected by consumer devices along with patient-reported symptoms to monitor the progression of respiratory and influenza-like illnesses." |
| Tavabi et al. (2022) <sup>36</sup> | "Digital biomarkers are physiological and behavioral measures collected from participants through digital tools that can be used to explain, influence, or predict health-related outcomes." |
| van den Brink et al. (2022) <sup>37</sup> | "Wearable technologies, including smartphones and smartwatches, are increasingly utilized in the healthcare domain for the development of so-called digital biomarkers. This novel type of biomarker is characterized by being measured non-invasively, continuously, and under real-world conditions using digital technology, allowing for a more holistic and personal insight into someone's health. Therefore, digital biomarkers enable accessible health and behavioral feedback to the user and are particularly suited for driving the healthcare transition towards prevention, empowering people in the self-management of health and disease. Furthermore, digital biomarkers can provide users with more frequent and detailed contextual information and continuously update personal lifestyle recommendations." |
| Dillenseger et al. (2021) <sup>38</sup> | "... digital biomarkers—digital health technologies—to explain, influence and/or predict health-related outcomes. Digital biomarkers stem is quite broad, and range from wearables that collect patients' activity during digitalized functional tests to digitalized diagnostic procedures and software-supported magnetic resonance imaging evaluation. With the increasing digitalization of healthcare, medicine now gains access to a new type of biomarker. So-called digital biomarkers enable the translation of up-to-date new data sources into informative, actionable knowledge. Digital biomarkers are basically collected by digital tools. Digital biomarkers mean objective, quantifiable physiological and behavioral data that are measured and collected by digital devices. The data collected by, e.g., |
|  | portables, wearables, implantables or digestibles are typically used to generate, influence and/or predict health-related outcomes, and thus represent deep digital phenotyping, collecting clinically meaningful and objective digital data.” |
| Hartl et al. (2021) <sup>39</sup> | “Digital biomarkers: Physiological and behavioral measures collected by means of digital devices such as portables, wearables, implantables, or digestibles that characterize, influence, or predict health-related outcomes.” |
| Gielis et al. (2021) <sup>40</sup> | “Complementary to their biological counterparts, digital biomarkers are “user-generated physiological and behavioral measures collected through connected digital devices to explain, influence and/or predict health-related outcomes.” |
| Hartl et al. (2021) <sup>39</sup> | “Digital biomarkers are defined as physiological and behavioral measures collected via digital devices (such as portables, wearables, implantables and digestibles) that characterize, influence, or predict health-related outcomes. Digital biomarkers offer several potential advantages compared to traditional clinical assessments. Digital biomarker products are usually the result of the combination of multiple individual hardware (sensors) and software (operating systems and algorithms) components. Digital biomarkers as clinical endpoints provide objective and quantitative measures yet still require broader clinical use and health authority acceptance.” |
| Sahandi Far et al. (2021) <sup>41</sup> | “Digital biomarkers (DB), as captured using sensors embedded in modern smart devices, are a promising technology for home-based sign and symptom monitoring in Parkinson disease (PD). The emergence of new technologies has led to a variety of sensors (ie, acceleration, gyroscope, GPS, etc) embedded in smart devices for daily use (ie, smartphone, smartwatch). Such sensor data, alongside other digital information recorded passively or when executing prespecified tasks, may provide valuable insight into health-related information. Such applications are now commonly referred to as digital biomarkers (DB). DB being collected frequently over a long period of time can provide an objective, ecologically valid, and more detailed understanding of the inter- and intra-individual variability in disease manifestation in daily life.” |
| Palanica et al. (2020) <sup>42</sup> | “Digital biomarkers are digitally collected data, such as heart rate from a wearable device, that are transformed through mathematical models into indicators of health outcomes like prediabetes. Some digital biomarkers have been found to outperform traditional clinical methods, for example, for arrhythmia detection, because of their ability to continuously monitor patients outside of the clinic. The most successful digital biomarkers have been developed based on supervised, unsupervised, and semi-supervised machine learning models.” |
| Babrak et al. (2019) <sup>6</sup> | “Digital biomarkers are objective, quantifiable, physiological, and behavioral measures that are collected by means of digital devices that are portable, wearable, implantable, or digestible. These data are often used to explain, influence, and/or predict health-related outcomes. Digital biomarkers fall within the scope of traditional biomarkers in relation to addressing health related questions, with use of a digital and portable technology that adds new dimensions, unique features, and challenges. digital biomarkers are usually less or non-invasive, modular, and often cheaper to measure. They can produce qualitative and quantitative measurements, but most importantly, they provide easier and cheaper access to continuous and longitudinal measurements.” |
| Nam et al. (2019) <sup>28</sup> | “In terms of IoT, the digital biomarker represents digitized data acquired from patients via IoT devices. Therefore, the digital biomarker can be defined as a biomarker that is objectively and quantitatively measured using digital devices and be used to explain or predict health-related outcomes. Digital biomarker is measured using the digital tools that include portable, wearable, implantable or digestible devices, and exclude data obtained via patient-reported measurements or traditional devices and equipment. In a broad sense, digital biomarker include all human data that can be measured using digital tool.” |
| Petersen et al. (2019) <sup>43</sup> | “The use of remotely collected data that monitors health and behavior is an emerging area of research. Such data could be considered digital biomarkers objective information that can be used to predict changes in health status and the use of digital biomarkers offers a more efficient method of identifying such markers as the use of devices continuously collecting data increases. One critical requirement in the development of digital biomarkers is connecting these novel measurements to health outcomes.” |
| Piau, Rumeau et al. (2019) <sup>44</sup> | “Digital biomarker definition. Objective, quantifiable, physiological, and/or behavioral data that are collected and measured by means of digital devices such as embedded environmental sensors, portables, wearables, implantables, or digestibles, and which opens up opportunities for the remote collection and processing of ecologically valid, real-life, continuous, long-term, health-related data.” |
| Seyhan et a. (2019) <sup>45</sup> | “Digital biomarkers (BMs) can have several applications beyond clinical trials in diagnostics—to identify patients affected by a disease or to guide treatment. Digital BMs present a big opportunity to measure clinical endpoints in a remote, objective, and unbiased manner. Digital BMs are defined as an objective, quantifiable physiological and behavioral data that are collected and measured by means of digital devices. The data collected is typically used to explain, influence and/or predict health-related outcomes.” |
| Zetterström et al. (2019) <sup>46</sup> | “We define a DB as patient-generated physiological and behavioural measures collected through sensors and other connected digital tools that can be used to monitor, predict and/or influence health-related outcomes.” |
| Dorsey et al. (2017) <sup>9</sup> | “Digital biomarkers – the use of a biosensor to collect objective data on a biological (e.g., blood glucose, serum sodium), anatomical (e.g., mole size), or physiological (e.g., heart rate, blood pressure) parameter followed by the use of algorithms to transform these data into interpretable outcome measures can help address many of the shortcomings in current measures. These new measures, which include portable (e.g., smartphones), wearable, and implantable devices, are by their nature largely independent of raters. They are, therefore, not prone to rater bias. The goal of digital biomarkers is to maximize the ecological validity and temporal and spatial resolution of capturing motor and nonmotor phenomena that are expected to change over time.” |
| Phillips et al. (2017) <sup>30</sup> | “Digital biomarker technologies, which fall into the category of “wearables and biosensing devices”, use consumer-generated physiological and behavioral measures collected through connected digital tools that can be used to explain, influence, and/or predict health-related outcomes. These |
|  | technologies may focus on measurements for consumer use only, or clinical measurements that are transmitted to clinicians for health care decision-making. They may passively monitor ongoing activities (such as steps taken) or be used to actively collect specific measurements (such as blood glucose).” |

## DISCUSSION

The first definition of a digital biomarker is from 2015 ^22^. Within eight years, more than 127 definitions have been used, with none of them clearly being the most widely used. These definitions often cover different aspects of definitional components that are traditionally used to describe more conventional biomarkers. Authors have created their own concepts and gave an identity to this type of biomarker. The variation in these definitions and the fact that only 23 of them provide a full description containing all relevant components, shows how broad the current understanding of this fundamental concept is.

Digital biomarkers emerged as a concept in medical and technological domains, albeit with a diverse terminology across different academic journals. In the medical field, digital biomarkers are often referred to as biomarkers of health or disease obtained through digital health technologies. In the technical field, these biomarkers are viewed as data-driven indicators collected from sensors, wearables, and other portable digital technologies that provide an assessment of the health status. These diverse terminologies and definitions reflect the interdisciplinary nature of digital biomarkers with their application in a broad spectrum of biomedicine which underlines the importance of unified concepts to enhance the communications and cross-disciplinary collaborations on this evolving field.

### Regulatory Perspectives

The EMA has defined digital biomarkers in 2020 in their draft guidance “Questions and answers: Qualification of digital technology-based methodologies to support approval of medicinal products”, stating their “clinical meaning is established by a reliable relationship to an existing, validated endpoint” ^10^. EMA draws a clear line to electronic clinical outcome assessments (eCOA), whose “clinical meaning is established de novo”. According to EMA’s terminology, both digital biomarkers and eCOA are derived from “digital measures” and can be used as “digital endpoints” ^10^.

On the other hand, the term “digital biomarker” cannot be found in the FDA draft guidance “Digital Health Technologies for Remote Data Acquisition in Clinical Investigations”, which instead features eCOA as examples of digital health technologies ^23^. Figure 3 contains our semantic interpretation of the terminology used by EMA and FDA.

**Figure 3.**
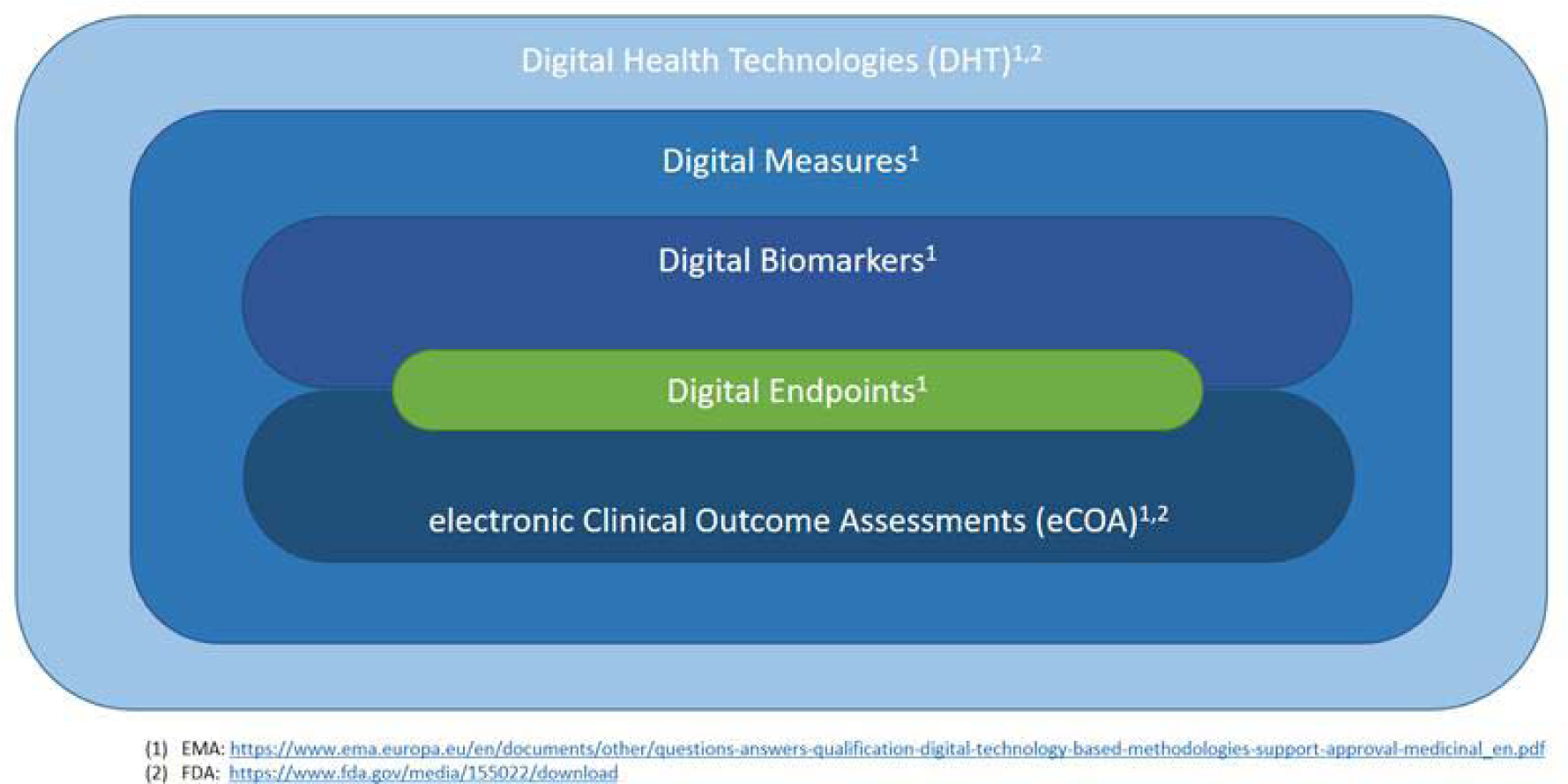
Semantic overview of terminology used by EMA and FDA Digital Health Technologies obtain Digital Measures which include Digital Biomarkers and electronic Clinical Outcome Assessments (eCOA). Digital Biomarkers and eCOAs both can provide Digital Endpoints.

This distinction can rarely be observed in the medical literature – we found this term in 8 of the 415 articles analyzed and a PubMed search for “electronic clinical outcome assessment*” returned also only 8 articles mentioning it in title or abstract (as of August 31, 2023), compared to the 415 for our search term “digital biomarker*”. As Vasudevan et al. stated in 2022: “There are currently multiple definitions of the term digital biomarker reported in the scientific literature, and some seem to conflate established definitions of a biomarker and a clinical outcomes assessment (COA)” ^11^.

This divergency in the terminology of digital biomarkers between the academic literature and the regulators’ language raises challenges and ambiguity. Consequently, a more cohesive and comprehensive framework within the digital biomarker field is needed to strengthen the clarity and continue growing the potential that this data could bring for health.

To achieve a common and more unified understanding of what digital biomarkers are – and are not – a Delphi approach could be useful ^24^. Such a study would aim to combine multiple views and expectations on the definition of digital biomarkers until a consensus is reached. Ideally, that would be achieved by an international panel with expert’s representative of all relevant stakeholders covering a range of medical fields (e.g., cardiology, neurology, etc.), professional backgrounds (e.g., clinical care/rehabilitation/nursing, software developers, device manufacturer, editors, guideline developers), and professional perspectives (e.g., academia, regulatory, industry/technology, publishing) and involving patients.

### Limitations

There are some limitations to our study. First, we used a limited search only in a single database using the single term of “digital biomarker*”, which may have overlooked some other relevant studies. It is very unlikely that the definitions would be much more uniform in these overlooked studies, and it is quite possible that many more different definitions would emerge. Therefore, we may have even underestimated the large number of different definitions. Second, the screening and data extraction was performed by a single reviewer only. This may have resulted in some studies that were overlooked and some misclassifications, but it is unlikely that our main interpretation would change. Third, we developed a simple framework with three key elements of definitions based on a well-established framework (BEST), but the categorization of elements is subjective to some degree. However, we employed a structured analysis that confirmed the observed heterogeneity across definitions.

## CONCLUSIONS

Clear and unambiguous communication and research reporting is essential for the effective implementation of scientific innovations and developments. This requires clear definitions and consistent use and understanding of key terms and concepts. A lack of clarity and consistency can lead to research waste, delay or even misdirection of promising developments and potential. Digital biomarkers offer the opportunity to collect objective, meaningful, patient-relevant data cost-effectively with unprecedented granularity. An exact understanding of what they are and how they are described in biomedical literature is essential to let them shape the future of clinical research and enable them to provide most useful evidence for research and care. Our study can inform the development of a harmonized and more widely accepted definition.

## Data Availability

All data that has been analyzed is provided in the supplementary material S2.

## SUPPLEMENTARY MATERIAL

**S1:**
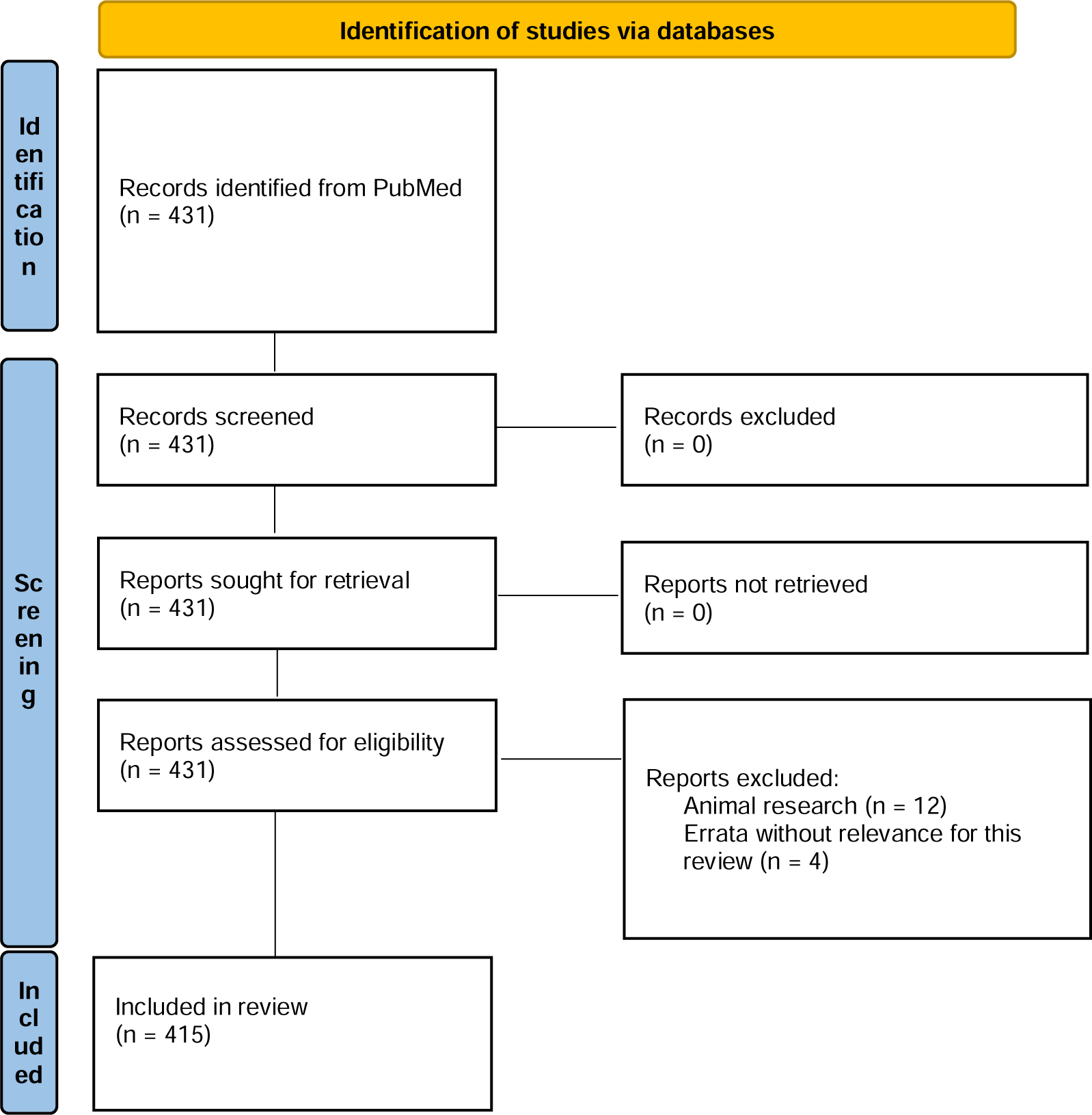
Flowchart illustrating the literature search and selection process

**S2.** Spreadsheet containing the (sheet 1) bibliography of all identified articles (n=415), (sheet 2) bibliography of all identified articles that provided a definition of digital biomarker (n=128), (sheet 3) characteristics of all identified definitions of digital biomarker (n=202), and (sheet 4) all unique identified definitions of digital biomarker (n=127)

**S3.**
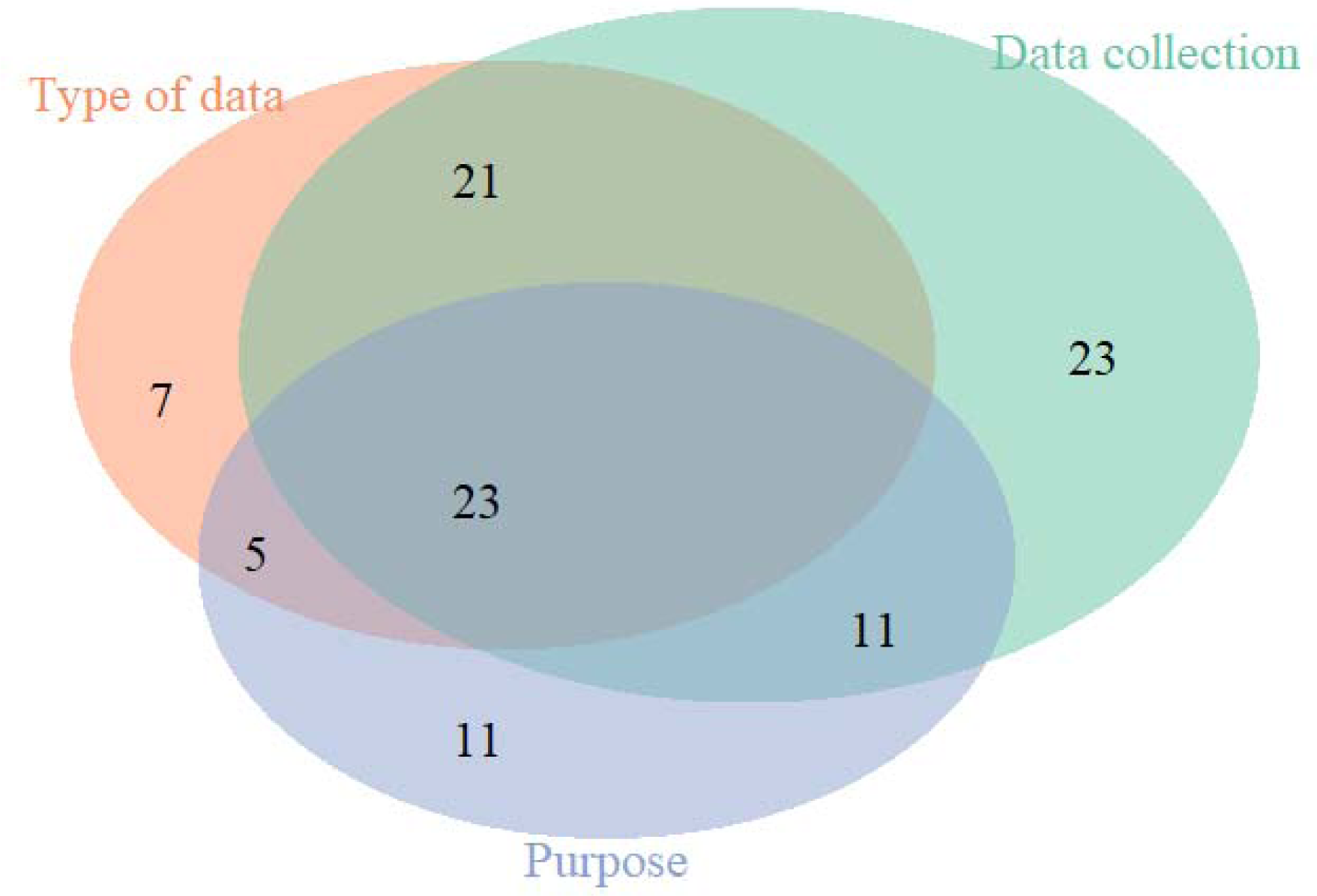
Venn diagram illustrating the components of the identified digital biomarker definitions (n=127)

**S3.**
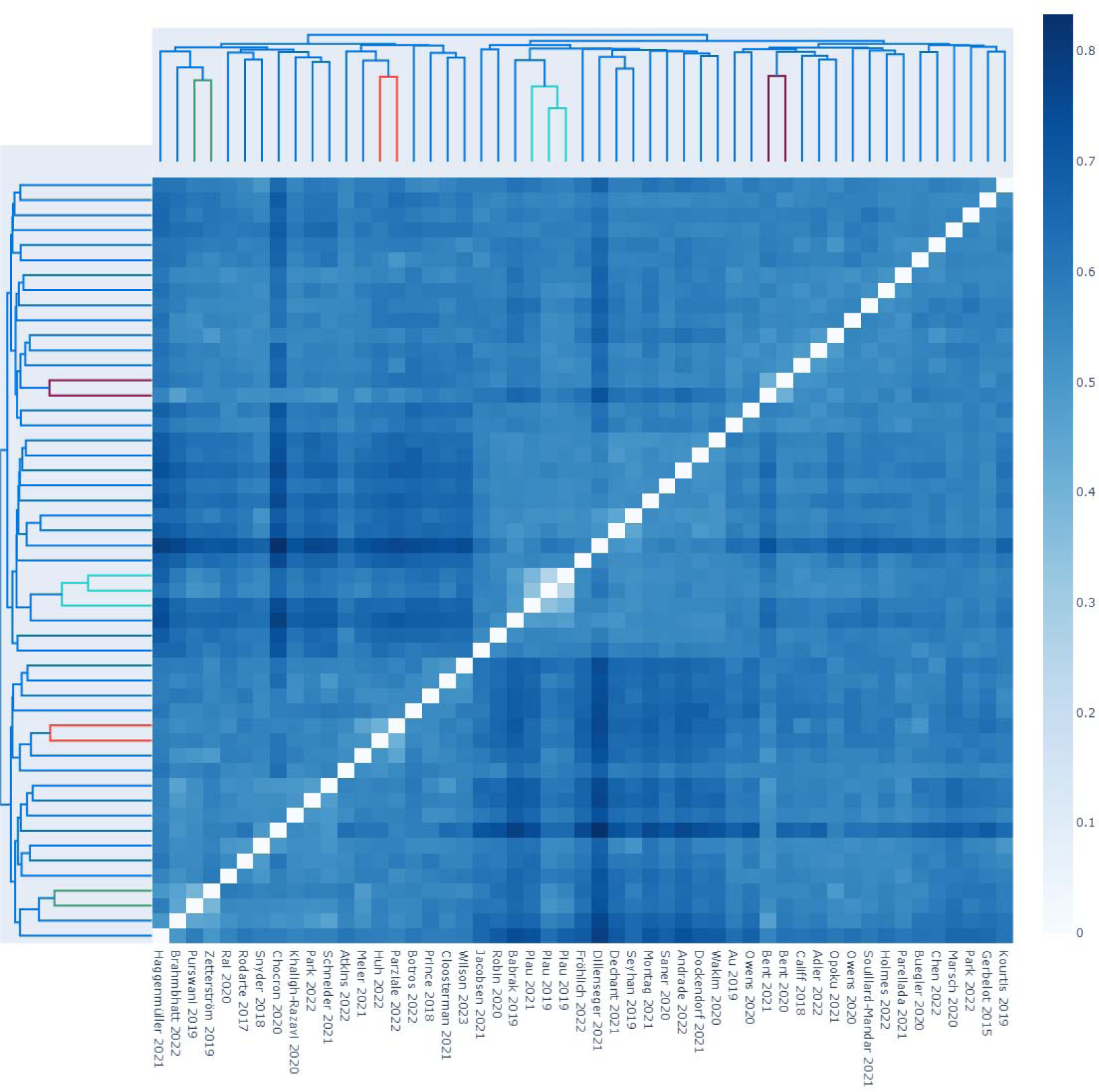
Symmetrical distance-matrix based on the structural Indel-distance of the 51 definitions without a reference (derived from 37 papers). A smaller distance (white / light blue) indicates structurally similar definitions, for which few insertions / deletions are required to change one definition into the other. A larger distance (dark blue) indicates structurally different definitions. The dendrograms at the top and left-hand side are derived through hierarchical-clustering and lead to more similar definitions being clustered next to each other.

## DECLARATIONS

### Ethics approval and consent to participate

Not applicable.

### Consent for publication

Not applicable.

### Availability of data and materials

All data that has been analyzed is provided in the supplementary material S2.

### Authors’ contributions

All authors made substantial contributions to the conception and design of the work; all authors have drafted the work or substantively revised it; all authors have approved the submitted version; all authors have agreed both to be personally accountable for the author’s own contributions and to ensure that questions related to the accuracy or integrity of any part of the work, even ones in which the author was not personally involved, are appropriately investigated, resolved, and the resolution documented in the literature.

## Acknowledgements

We thank Saido Haji Abukar (Medical Bachelor student at ETH Zurich) for her support with the study selection and data extraction.

